# The Power of Touch: How can Social Touch impact the Trust, Body Representation and Exercise Performance of Patients in a Wearable Physiotherapy Assistant?

**DOI:** 10.1101/2025.05.16.25327433

**Authors:** Nicola Visentin, Merle Fairhurst, Wenhan Sun, Xinyao Niu, Philipp Floessel, Freya Charlotte Wunderlich, Lisa-Marie Lüneburg, Stefan Teubner, Willy Beyer, Benas Sudzius, Doris Lachman, Hagen Malberg, Jens Krzywinski, Alexander Disch

## Abstract

**Background:** Given the high prevalence of low back pain and its financial weight on the healthcare system, practicing physiotherapy exercises is crucial for sustainable therapy success. Effective rehabilitation requires high-quality movement execution, demanding technology that provides optimal feedback.

**Objective:** Our interdisciplinary approach—combining neuroscientific insights on body representation, sports science findings on motor learning, and user experience research on feedback perception aims to give a multifaceted insight into the different effects that varied feedback modalities (specifically, auditory, haptic, and combined), have on the performance of a physiotherapy exercise, and on the cognitive workload and body image representation of the patients during the exercise itself, in people with and without nonspecific back pain.

**Methods:** This study employs a mixed-methods design to investigate the impact of different feedback modalities (auditory, haptic, and combined) on physiotherapy exercises. In a quantitative Wizard-of-Oz experiment (n=57), participants performed bent knee side planks while receiving feedback—secretly provided by physiotherapists—via a smart shirt. Outcome measures included cognitive workload (NASA TLX), body image representation (Body Map Task), and exercise improvement (physiotherapist evaluation). Control variables such as trust in technology (Surgical Robot Trust & Trust in Automation Questionnaires) and pain level (Chronic Pain Grade Scale) were also assessed. A semi-structured interview gathered qualitative insights into participants’ feedback perception and usability.

**Results:** Results indicate no significant differences in cognitive workload or body image representation across modalities, though qualitative data suggest a preference for haptic and combined feedback over auditory alone. Performance outcomes did not significantly vary across conditions, but qualitative insights highlight the benefits of multimodal feedback in enhancing movement perception and engagement.

**Conclusion:** Findings suggest no single optimal feedback modality, but combining haptic and auditory cues enhances usability and motor learning. Participants favor this approach, initially relying on auditory and haptic feedback long-term. Despite not clearly emerging from quantitative statistical analysis, these results support the development of a multisensory feedback strategy. Our multi-perspective approach demonstrates that multimodal feedback is not only beneficial but necessary for designing adaptive, accessible, and effective rehabilitation technologies.

## 1 Introduction

By 2035, despite increasing healthcare demands, nearly 40 percent of rural areas in Germany will remain medically underserved. Physiotherapy, essential for treating back pain affecting almost two-thirds of the population, accounts for over 80 million treatments annually, costing up to 10 billion euros yearly [1]. The associated economic losses due to work disability amount to 28.6 billion euros. Physiotherapy’s personnel-intensive nature reinforces the healthcare gap, necessitating digital solutions operating in real-time for comprehensive provision. Low back pain is one of the most prevalent musculoskeletal disorders worldwide, with a lifetime prevalence of 80 to 85% [1, 2]. The financial burden on healthcare systems is anticipated to rise further in the coming decades.[3, 4] Recent studies have shown that well-developed trunk stability is crucial for this purpose in the general population [1, 5, 6, 7]. Also, they have shown that trunk-stabilising sensorimotor training performed twice a week for twelve weeks can reduce recurring rates in LBP patients by 80% [8, 9]. Consequently, the majority of them are treated in physiotherapy practices [10, 11]. Most people use an outpatient rehabilitation facility for this purpose. However, physiotherapy treatment is one of the most staff-intensive areas of medical care. An alternative form of treatment that can take place in the rehabilitant’s familiar surroundings is home-based training. This approach has been shown to facilitate the integration of treatment into the patient’s daily routine. In this context, independent home practice is becoming increasingly important. It can reduce the costs of the same rehabilitation success by about 50% [12]. To ensure the effectiveness of new technologies supporting home-based rehabilitation, it is essential to ensure patient acceptance and trust. Feedback mechanisms must be designed to be easily understood by patients and motivating, thereby fostering long-term adherence to the training programs.

### 1.1 Background

Recent studies in the field of post-operative treatments indicate that independent but significantly intensified home practice can achieve comparable rehabilitation success to guided physiotherapy [13]. To ensure this, patients must integrate rehabilitation measures into their daily routines. However, the effectiveness of home-based training depends heavily on individual discipline, self-motivation, and the correct execution of exercises. The combination of professional physiotherapist-led treatment and independent therapy intensification at home is key. High-quality movement execution is crucial for successful rehabilitation. During professional treatments, this is often achieved through direct feedback from therapists, delivered via verbal communication or physical touch. This social-affective touch not only provides guidance on movement but also fosters trust and a sense of connection, which can enhance the patient’s engagement with the therapy. In the context of home-based training, where physiotherapists are not present, alternative strategies must be employed to maintain this essential movement quality and build trust in the therapeutic process. Emerging technologies, such as wearable sensors or interactive applications, can simulate elements of therapist feedback. These technologies should incorporate mechanisms replicating the motivational and trust-building aspects of social-affective touch, enabling patients to feel supported and confident in their rehabilitation journey.

### 1.2 The Tactile Internet: a new possibilities for real-time human-machine communication

The tactile internet introduces new technical possibilities for delivering real-time feedback during home-based rehabilitation. With its near latency-free data transmission, it enables precise measurement and almost instantaneous feedback delivery [14]. One promising application is a vibrotactile-controlled feedback system, which not only supports autonomous training but also monitors exercise quality and guides patients in performing movements correctly [15]. This tactile feedback, perceived through the skin, can be generated by pressure or vibration, the latter being known as vibrotactile feedback [16]. In professional physiotherapy, tactile feedback is often provided directly by therapists to correct and optimize movements. In a home-based, hands-off context, however, a human-machine interface becomes essential to deliver this feedback. Vibrotactile systems typically employ small, lightweight vibrating actuators, which can be positioned on various body parts and tailored to respond effectively to specific stimuli [17, 18, 19]. The VEIIO project explores a novel approach to this challenge, developing a haptic physiotherapy assistant in the form of a smart shirt [20]. This system combines vibration and auditory feedback to enable real-time, multimodal human-machine interaction. By providing feedback comparable to that of a physiotherapist, the VEIIO solution aims to deliver a comprehensive and effective 1-on-1 training experience at home, ensuring high movement quality and fostering trust in the therapy process.

### 1.3 Social Affective Touch

The social connection between patients and physiotherapists is crucial for the acceptance of these kinds of autonomous systems. Recent research indicates a link between social touch and trust [21]. Social touch typically includes a communicative exchange between two individuals [22]. However, with the advent of affective haptics, mediated touch to be a powerful tool to enhance social connection. By incorporating touch into a multisensory feedback context more similar to face-to-face interpersonal interactions, patients will experience improved therapy quality, enhanced understanding, responsibility, motivation, and engagement.

### 1.4 Research Questions

However, research, as well as the use of vibrotactile feedback devices in a therapeutic context, is still insufficient, so little can be said about the design of technical feedback systems [17]. The aim of this research project is to derive requirements for the design of a tactile feedback strategy, especially for the implementation of trunk-stabilising exercises. To this end, this study examines the feedback behaviour of healthy people and analyses their use of technical tactile feedback.

1. Cognitive workload: does the combined auditory and haptic feedback result in a different cognitive workload for participants compared to conditions where only one modality of feedback is provided?
2. Body image representation: do the haptic feedback condition and the combined feedback condition lead to better body representation compared to the auditory feedback condition?
3. Is the feedback given by the combined modality feedback better in terms of performance improvement compared to the only haptic and the only auditory feedback conditions?

## 2. Materials and Methods

### 2.1 Sample

The sample consisted of 29 women, 27 men and one non-binary with an average age of 28.9 years (SD*±*10.5), an average height of 174.4 cm (SD*±*10.3), and an average weight of 70.5 kg (SD*±*11.8) [Table A.1 SM]. Out of these 57, 8 participants showed chronic back pain, as per the Chronic Pain Grade Scale (see also section 2.2). The research project was approved by the Ethics Committee of Technische Universität Dresden (BO-EK-215052022) and follows the Declaration of Helsinki. At the outset of the study, all participants provided written informed consent, which included comprehensive details about the study and the privacy policy (data processing by video recording). Participants received a compensation of 15 € for the work time lost due to their involvement in the study.

### 2.2 Methods

This study employed a mixed-methods approach, combining quantitative data from a Wizard-of-Oz experiment (WoZ) with qualitative insights from post-experiment interviews [23]. Within the WoZ experimental setup a physiotherapist manually provided randomized feedback modalities for motion correction by a smart shirt (described in section 2.3). During the experiment, participants experienced three feedback modalities performing a bent knee side plank exercise:

- **Haptic feedback** (delivered via the smart shirt)
- **Auditory feedback** (delivered through PC speakers)
- **Combined feedback** (haptic and auditory feedback simultaneously)

The goal of the study is to compare the effectiveness of the three feedback modalities by following **dependent variables**:

- **Workload on participants**, measured with the NASA Task Load Index (NASA-TLX).
- **Body image representation**, measured with the Body Map Task (BMT) (code at github), in which participants were presented with 2D avatars (front and back) and instructed to use a palette of colour shades from turquoise to red to paint different parts of the body to create heatmaps indicating body regions that they felt the most activated during the exercise.
- **Improvement in the performance**, measured with the Physiotherapist evaluation form for correction behaviour on 4-point Likert scale, included 3 items: adjustment based on feedback; alignment with feedback; alignment without feedback
- **Feedback usefulness**, measured by the number of feedbacks that were given out during each exercise

Our independent variable is the modality of feedback. Furthermore, the study included the following as **control variables**:

1. **Pain level during the previous 3 months**, measured by the Chronic Pain Grade Scale (CPGS) [24]; this also served to categorize the participants in two different groups (Pain vs No pain) for the qualitative interviews performed at the end of the session
2. **Trust in technology**, measured by the adapted questionnaires of Trust in Automation (TIA) [25] and the Surgical Robot Trust Questionnaire (SRTQ) [26]
3. **Demographic variables**: age, gender, weight, and height
4. **Number of repetitions** during each block

Data collection was done using the Gorilla software, which also randomized the different conditions; data analysis was then conducted using the statistical software RStudio. For the qualitative analysis of the semi-structured interviews, we used the software MaxQDA. The interviews were audio-recorded and transcribed using the integrated tool by the same software. The study procedure, illustrated in Figure X, consisted of three phases:

1. Pre-condition data acquisition using validated questionnaires.
2. The Wizard of Oz (WoZ) experiment with three randomized feedback conditions.
3. Post-condition data acquisition involving adapted questionnaires and a semi-structured interview focusing on trust and workload.

At the beginning of each session, participants were informed about the study goals and were told that the smart shirt uses AI to provide real-time movement corrections. The pre-condition phase included demographic questions and the measurement of the pain levels during the last 3 months by the CPGS. Following this, participants wore the smart shirt and familiarized themselves with the three feedback types. After receiving instructions on the exercise from the physiotherapist (PT), the WoZ consisted of **three randomized blocks**, one for each feedback modality. Each block included:

1. Instructions about the upcoming feedback modality.
2. 10-15 repetitions of the exercise, during which also the number of feedback given was counted; for the exercise, participants were instructed to perform a bent-knee side plank exercise at a consistent difficulty level (see **Figures** below). The PT provided a detailed explanation of the exercise, beginning with the correct starting position and the intended movement pattern. Common movement errors were discussed in relation to the feedback that had been previously assessed. The PT then guided the participants through the process of recognizing and responding to the various types of feedback, both auditory and haptic, while offering specific instructions on how to adjust their movements in response to the feedback or avoid common errors.
3. Post-exercise measure: BMT, NASA-TLX, Physiotherapist evaluation form for correction behaviour on 4-point Likert scale

After the final block, post condition was acquired by completion of several questionnaires and a semi-structured interview:

- Second part of the NASA-TLX, for dimension weighting.
- SRTQ and TIA
- Semi-structured interview: a guide with four open-ended questions was developed with the physiotherapist to qualitatively evaluate the participant’s experience and explore factors influencing the feedback perception. Interviews took place in person after all interviews were audio-recorded and transcribed. All data were collected by the same interviewer, held and analyzed in accordance with the Data Protection Act, with the actual data available only to the authors of the present paper.

The study was conducted with both German-speaking and English-speaking participants; therefore, for each questionnaire and instrument involved two versions were used, one for each language. For the questionnaires where only one of the two versions was present and validated, a translated version was created by the researchers: this concerns the CPGS, the BMTs instructions, and the SRTQ. The latter was therefore included in the study only as an exploratory variable since it lacks statistical validation.

**Figure 1.**
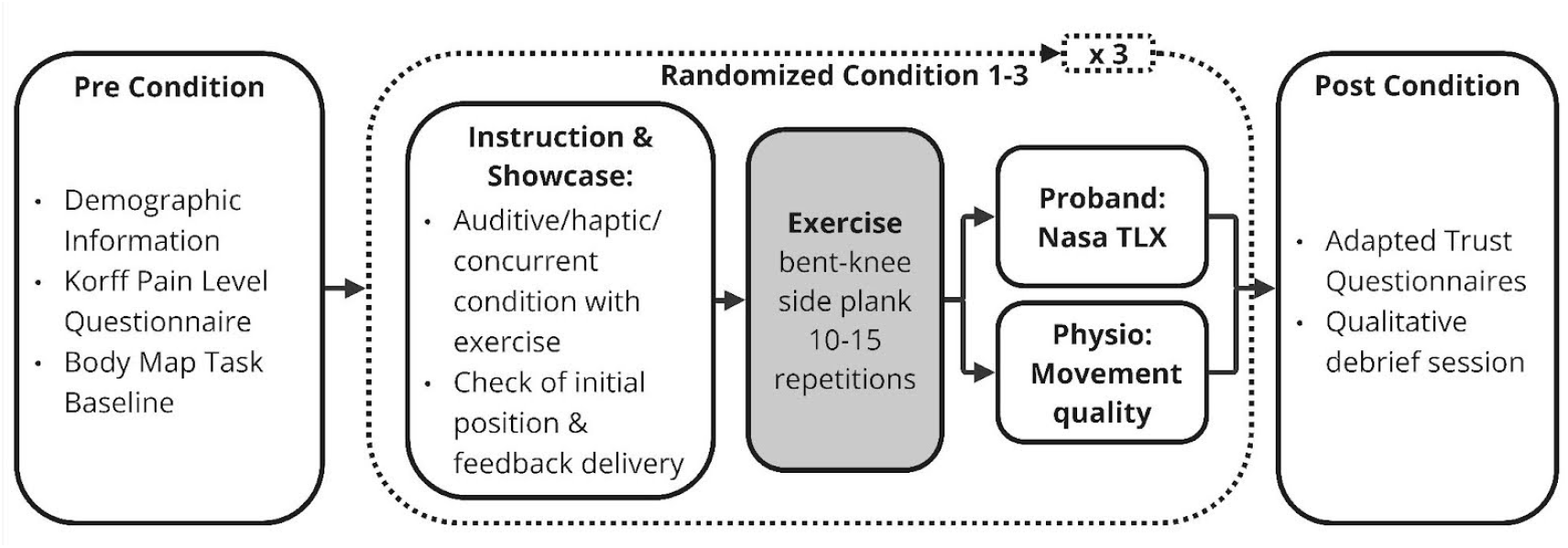
Flowchart of the study.

### 2.3 Technical Implementation of the Smart Shirt

The smart shirt was conceptually designed to integrate motion-detection sensors with actuators to provide corrective vibrotactile and auditory feedback for movement errors. While presented as automated in our WoZ experiment, the system in this study delivered feedback manually via a graphical user interface (GUI) operated by physiotherapists, allowing realistic simulation without automation complexity. Therefore, the test subjects wore a tight-fitting long-sleeved top containing 9 DC vibration motors providing vibration feed-back. The system’s architecture ensured minimal feedback delivery delay, averaging approximately 50 ms, providing a timely and realistic experience. Vibrotactile signals for the haptic feedback were delivered using embedded vibration motors. Patterns involved one or multiple actuators activated simultaneously at approximately 50 Hz and full intensity for 1000 ms or 2000 ms. Placement and intensity were optimized for clear perception and interpretation. Auditory feedback, triggered via the GUI, consisted of pre-recorded instructions delivered through external speakers. Instructions were provided in German or English, depending on participant preference, ensuring clarity and consistency (Duration 3000 ms 4000 ms). The smart shirt deployed auditory, haptic or both feedback modalities combined to correct common movement errors in the upper body and core during training.

All shirt components are insulated and shielded from the test subject by housings. The applied voltage of 5 V and a current of 0.9 A pose no danger to healthy test subjects in the event of a defect. The presentation of the exercises took place in a laboratory in the area of the chair of industrial design engineering. The physiotherapists were in the same room as the subjects while they were correcting them via different feedback modalities. During the exercise performance, the physiotherapist also gave no other feedback. The subjects could concentrate fully on the technical feedback while performing the exercise. The physiotherapists were experienced in using the devices and providing feedback. The therapist was significantly involved in the six-month development phase.

**Figure 2.**
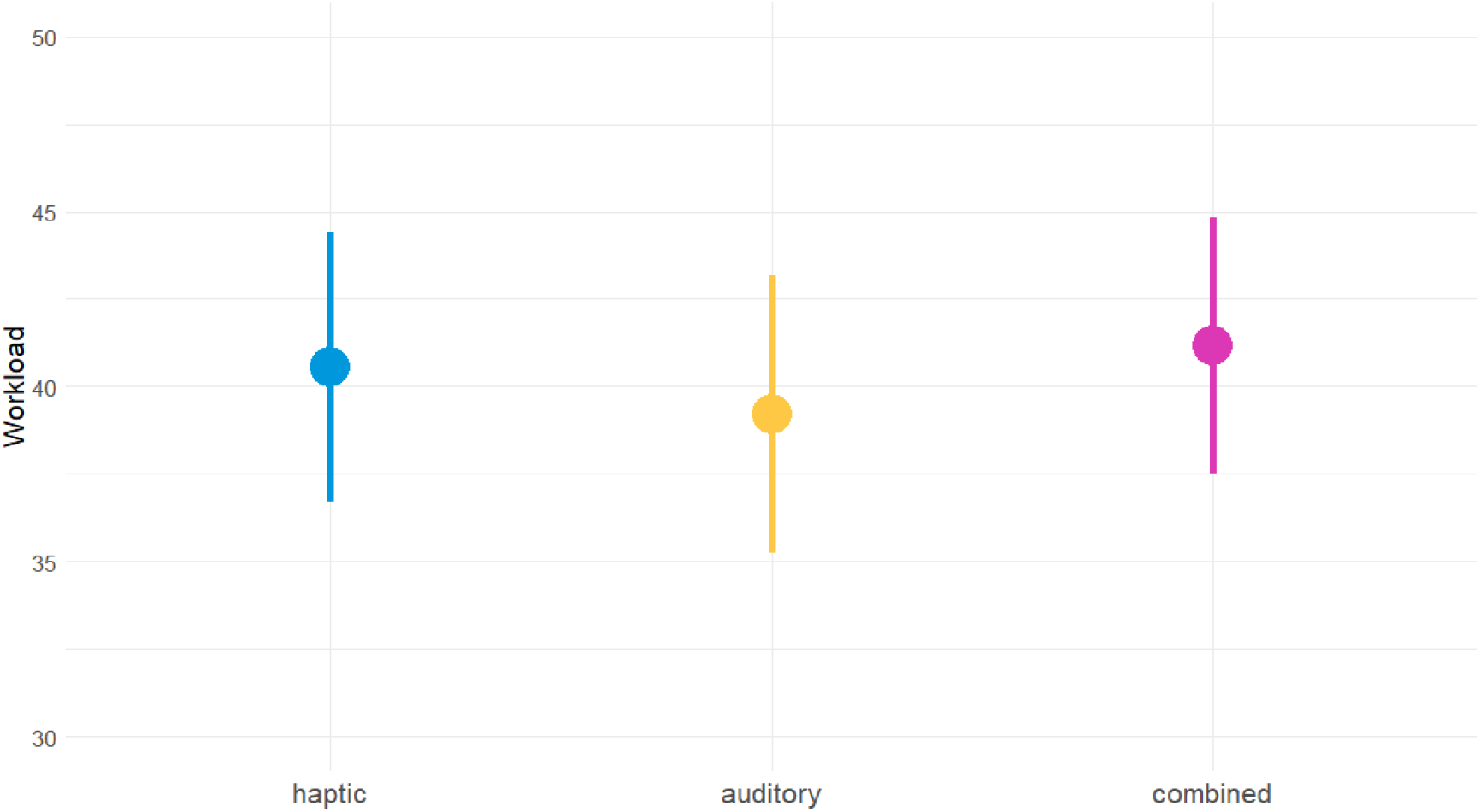
Mean workload on participants, measured via NASA-TLX, in different feedback modalities.

## 3. Results

### 3.1 Workload on Participants

Using an ANCOVA, controlling for users’ trust in technology, as well as for demographic variables such as gender, age, height and weight, the statistical analysis revealed the main effect of the feedback modality (auditive, haptic, combined) to be non-statistically significant, and very small (*(F(2, 145) = 0.323, p=0.725)*); taking into account the various control variables included in the study design, the ones that showed to have a significant effect on the workload are participants’ height (small effect, *F(1, 145) = 8.10, p = 0.005*), and two subscales of the TIA questionnaire: Intention of Developers (medium effect, (*F(1, 145) = 7.34, p = 0.008*), and Familiarity (small effect, *F(1*,*145)=5.31, p=0.023*) [Table A.2 SM]. These results show that the presence of a haptic feedback, carried out by the feedback shirt, does not have an influence on the participants’ workload, as the differences between groups can be explained mainly by people’s perceptions of technology in general [Figure 4].

**Figure 3.**
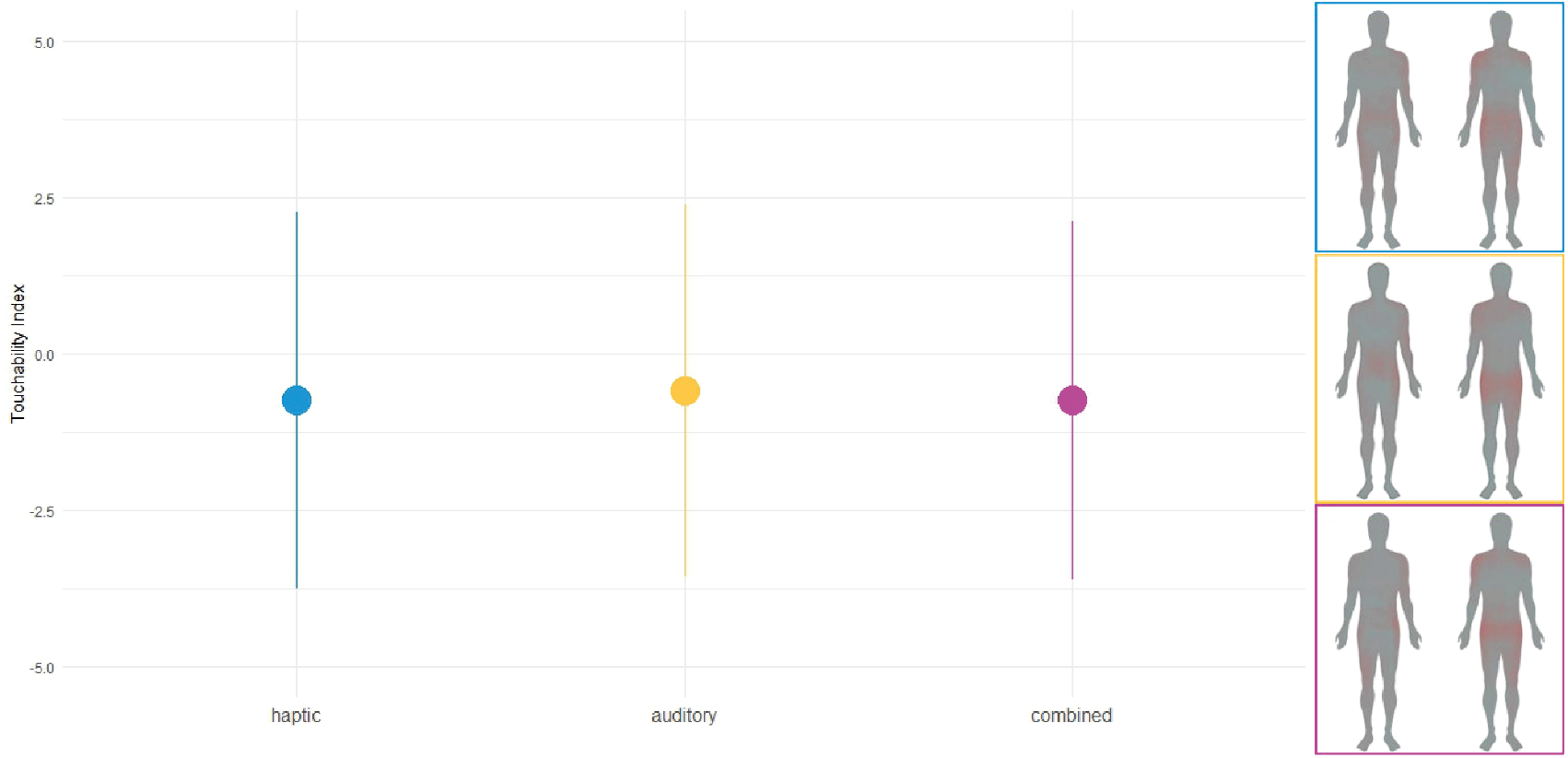
Mean touchability index in different feedback modalities. On the right, representation of the mean heatmap colored by participants in the Body Map Task for different feedback modalities with adapted visual: brightness increased 20%; contrast increased 30% for better visibility.

**Figure 4.**
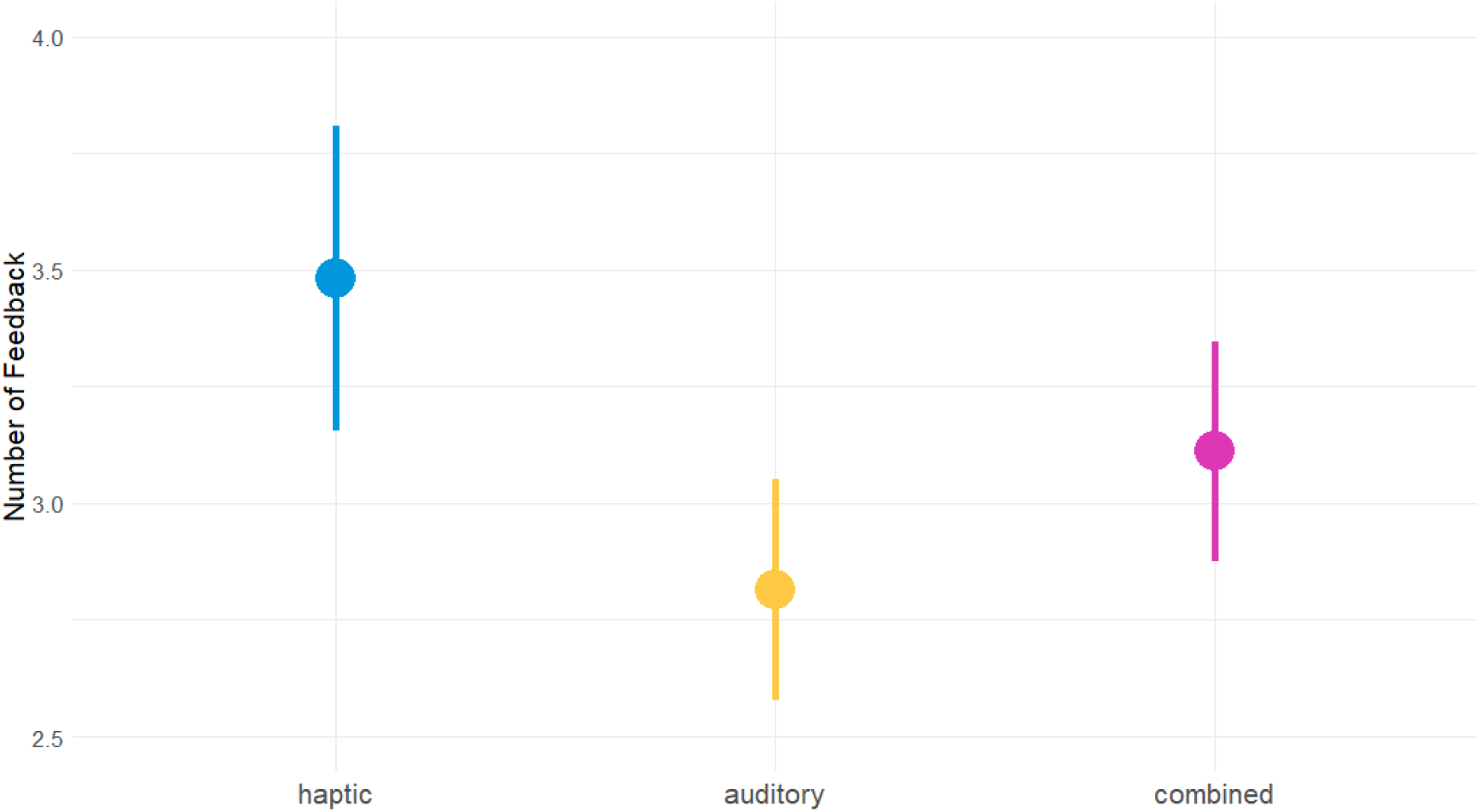
Mean number of feedback given out during the exercise in different feedback modalities.

### 3.2 Body Image Representation

For the BIR, participants were asked to fill out a BMT after each one of the three blocks. Then, an ANOVA was carried out to determine the different impact of different feedback modalities on the BIR. Results show that the main effect of modality is statistically not significant and very small (*F(2, 681) = 0.01, p = 0.988; η*^2^ = 3.57*e* − 05, 95% *CI [0.00, 1.00*]) [Table A.3 SM].

### 3.3 Improvement in the performance

To measure the improvement in the performance of the participants, the physiotherapist filled out, after each block, the Physiotherapist evaluation form for correction behaviour on 4-point Likert scale. The questionnaire’s first three items were considered for this variable, while the other three were used to measure the feedback usefulness (see 3.4). Of the 57 initial participants, 3 of them were removed from the final analysis of the third item due to missing data. For each one of the three items an ANOVA was carried out to determine the impact of the feedback modality on it. For all three items the main effect of the feedback modality is statistically not significant: very small for the first item (*F(2, 168) = 0.58, p = 0.561; η*^2^ = 6.85*e* − 03, 95% *CI [0.00, 1.00]*), and small for the second (*F(2, 168) = 1.31, p = 0.271; η*^2^ = 0.02, *95% CI [0.00, 1.00]*) and third (*F(2, 159) = 0.94, p = 0.391; η*^2^ = 0.01, 95% *CI [0.00, 1.00]*) [Table A.4 SM].

### 3.4 Feedback Usefulness

For the last dependent variable, the feedback usefulness, the last three items of the Physiotherapist evaluation form were taken into consideration. Of the 57 initial participants, 3 of them were removed from the final analysis due to missing data. The dependent variable on this part of the analysis was the number of feedback given out during each block, while the number of repetitions and the item 4 of the questionnaire were used as control variables. An ANCOVA was carried out to determine the effect of the feedback modality on the number of feedback, while also controlling for the trust in technology (SRTQ and TIA) and for demographic variables. The main effect of the feedback modality proved to be statistically significant and medium (*F(2, 114) = 8.33, p ¡* .*001; η*^2^(*partial*) = 0.13, *95% CI [0.04, 1.00]*). In this model, some of the control variables also showed significant influence on the number of feedback, specifically the number of repetitions (medium effect, *F(1, 114) = 13.00, p ¡* .*001;η*^2^(*partial*) = 0.10, *95% CI [0.03, 1.00]*), item 4 of the Physiotherapist evaluation form (large effect, *F(1, 114) = 27.85, p ¡.001; η*^2^(*partial*) = 0.20,*95% CI [0.10, 1.00]*), and two of the demographic variables, specifically age (small effect, *F(1, 114) = 5.85, p = 0.017;η*^2^(*partial*) = 0.05, *95% CI [4.77e-03, 1.00]*), and weight (large effect, *F(30, 114) = 1.96, p = 0.006; η*^2^(*partial*) = 0.34, *95% CI [0.06, 1.00]*)[Table A.5 SM]. Post hoc comparison using the test with Bonferroni correction indicated that the mean score for the haptic modality (*M=3.481, SD=1.255*) was significantly different than the auditory modality (*M=2.814, SD=0.913*). However, the combined modality (*M=3.111, SD=0.904*) did not significantly differ from the auditory and the haptic modalities [Table A.6 SM].

### 3.5 Qualitative Study Data Analysis

The data analysis of the qualitative interviews provided in-depth insights into the factors influencing participants’ workload, body perception, and the perceived usefulness of the feedback modalities. Therefore, a structuring content analysis and a coding system were created [27]. All questions, responses, and quotes were translated from German, with some exceptions, such as when the interview was in English. A qualitative content analysis, as outlined by Kuckartz [28], was employed. Ten interviews—equally split between healthy individuals and back pain patients—were randomly selected for analysis. A category system was developed deductively and refined through trial coding with three interviews, during which subcategories were added inductively. Two coders independently analyzed the remaining data, resolving disagreements through discussion and review with a third team member.

### Haptic feedback

The majority of participants described the haptic feedback as clear, effective, and pleasant, with an emphasis on the clarity and directness of the vibration signals. Some, however, associated the vibrations with stressful stimuli.

> *“(So) I found the haptics much more intuitive. The body also simply understood it more quickly.” (Proband A)*

*Workload and perceived usefulness:* Haptic feedback was found to be intuitive and easy to interpret after a short adaptation period. Participants without prior experience initially had to concentrate more to assign the vibrations to specific movement corrections correctly.

> *“(But) sometimes, I had to really focus and think about where it was vibrating exactly.” (Proband B)*

#### Body perception

The participants’ perceptions of intensity were strongly influenced by the quality of contact between the actuators and their skin. They noted that direct contact produced clearly noticeable yet comfortable feedback, and most agreed that the actuator positions were logical.

#### Challenges in Haptic-Only Feedback

Participants highlighted several challenges associated with using haptic feedback as the sole modality during the exercises. These challenges primarily revolved around the initial interpretation of the signals, physical factors of the smart textile affecting the feedback perception, and individual preferences. It was especially challenging for participants with no prior exercise experience, as they were unfamiliar with associating specific vibrations with corrections.

### Auditory feedback

Participants viewed auditory feedback as a valuable supplementary input during initial use and generally perceived it as intuitive and effective. While some participants favored it as much as haptic feedback, others found it confusing or irritating.

#### Workload and perceived usefulness

A few participants noted that auditory feedback was easier to implement compared to haptic signals, yet many reported that it was more confusing and exhausting to process, often requiring additional time to interpret the cues. They reported needing additional time to interpret auditory cues, which occasionally disrupted their focus and form. They reported needing additional time to interpret auditory cues, which occasionally disrupted their focus and form.

> *“I found the purely auditory feedback a bit confusing. [*… *] Without the haptics, it was noticeably harder because you take a moment to listen and think, ‘Okay, let’s see, what did that actually mean?” (Proband A)*

*Body perception:* Some participants preferred the brief and concise instructions, as they were able to respond to them quickly and effectively. Furthermore, auditory feedback was sometimes perceived as more natural, and participants found it easier to perceive their body in response to this feedback modality. Some participants found it more difficult to perceive their body through the auditory cues alone, as the feedback did not directly correspond to the body but had to be processed through the auditory sense first.

#### Challenges with Auditory-Only Feedback

The duration and therefore occuring delay in understanding auditory feedback led to moments of hesitation, where participants had to “search” for the intended signal within their body. This process sometimes created a feedback loop of errors, making it harder to maintain proper execution of tasks.

> *“Because then you start feeling around in your body, and you might even neglect your form, potentially getting into a little downward spiral. [*…*] You lose focus, then think, ‘Oh wait*,*’ and end up receiving more and more feedback.” (Proband A)*

### Combination of feedback conditions

The combination of haptic and auditory feedback was well-received and seen as complementary. Participants reported that using both modalities together provided a sense of security and improved the speed and clarity of interpreting feedback, particularly when haptic signals were weakened by misalignment or poor skin contact. Although both modalities were valued individually, their combination was generally preferred for its enhanced clarity during initial use.

*Workload and perceived usefulness:* Participants found the dual-modality approach especially beneficial in the early stages of therapy or training, as it facilitated quicker adaptation to the feedback system by reducing ambiguities associated with using a single modality. Over time, as users became more familiar with the haptic cues, the reliance on auditory feedback diminished.

> *“Well, I think the vibration is good. I just think that, initially, for a certain period during therapy or training, you still need the auditory cues, but after a while, once you’ve gotten used to it, they’re no longer necessary.” (Proband C)*

*Body perception:* The combination was described as a robust solution for scenarios where precise feedback was critical, particularly in overcoming the limitations of a single modality. This approach provided flexibility, catering to individual preferences and ensuring accessibility for a wider range of users.

## 4. Discussion

The objective of this study was to examine the feedback behaviour of healthy individuals and those with unspecific back pain conditions, comparing tactile feedback (vibrations), auditory feedback, and the combined condition. Its particular strength lies in its mixed-methods design, combining quantitative data with qualitative insights, thereby providing a comprehensive understanding of how different feedback modalities impact both performance and patient experience in digitally assisted physiotherapy exercises. The study examined whether combined auditory and haptic feedback increases cognitive workload, improves body representation, enhances performance, and investigates the perceived usefulness of the different modalities.

### 4.1 Cognitive Workload

The results show no statistically significant differences in cognitive workload across the different feedback conditions, aligning with previous research that has investigated the impact of feedback modalities on task load [29]. Similarly, combining feedback modalities did not significantly alter participants’ perceived workload. In the context of robot-assisted tasks, research has emphasized the importance of multimodal sensory feedback in reducing cognitive load, enhancing user experience, and improving accessibility [30]. However, qualitative data revealed a clear preference for haptic and combined modalities over auditory feedback only, with participants finding haptic feedback easier to process and quicker to understand, whereas auditory feedback was more overwhelming. These findings support our earlier feasibility study [20], demonstrating that haptic feedback can enhance movement quality without increasing mental workload. Haptic feedback in upper limb motor therapy improves performance and reduces mental workload, making it more useful, easier to use, and less difficult to learn [31]. Interestingly, participants also found the combination of haptic and auditory feedback especially helpful in the early stages of training, adding faster adaption. This observation is consistent with research on multimodal interfaces, which shows that combining auditory, haptic, and visual feedback can not only enhance performance but also reduce mental workload [32]. Taken together, these findings highlight the potential of multimodal feedback systems in improving both user experience and performance without adding undue cognitive load.

### 4.2 Body Image Representation

The results of this study indicate no statistically significant differences in body image representation across the different feedback conditions, while qualitative data presents mixed findings. This aligns with prior research suggesting that proprioceptive body models remain stable across real and imagined conditions, implying the presence of a common, stored representation of the body’s metric properties that is resistant to short-term external modulation [33]. The inherent distortions in body representation may, therefore, persist despite variations in feedback modality, as the central nervous system relies on a stable internal body schema. However, proprioceptive acuity varies across different joint positions, suggesting that while proprioceptive feedback can influence body representation, it does not necessarily translate into observable changes in body image across different feedback conditions [34]. This may explain why no significant differences emerged in our quantitative data despite qualitative reports suggesting that multimodal feedback—particularly haptic feedback—enhanced participants’ perception of movement accuracy. The effectiveness of feedback highly dependent on the physical adherence of actuators to the skin, which could impact the fidelity of sensory input and the way it is processed by the brain. Neuroscientific evidence suggests that body image representation is shaped by the integration of visual, tactile, and proprioceptive information in key cortical regions such as the posterior parietal cortex and premotor cortex, which are crucial for updating limb position and maintaining body ownership [35, 36]. The role of these regions highlights the complexity of sensory integration mechanisms, where multisensory inputs contribute to a more precise body schema but do not necessarily produce immediate, measurable changes in subjective body image representation. While our findings do not indicate significant alterations in body image representation across feedback conditions, they support the idea that proprioception remains the dominant modality in body schema stability, with multimodal integration playing a crucial role in refining movement perception. The mixed qualitative responses underscore the complexity of sensory integration, suggesting that individual differences and the nature of sensory input delivery may critically influence the perception of one’s own body during movement.

### 4.3 Improvement in the performance

The results indicate that haptic feedback alone does not significantly improve performance compared to auditory or combined feedback despite an initial period of adaptation. This finding is consistent with research suggesting that concurrent feedback can enhance immediate motor performance but may interfere with long-term motor learning, depending on the timing and purpose of the feedback [37]. To develop an effective feedback strategy, it is essential to align feedback timing with its intended goal, whether to enhance performance or facilitate motor learning. Additionally, the motor skill level of participants plays a crucial role, as concurrent feedback tends to be most beneficial for individuals with lower skill levels [38]. This highlights the necessity of evaluating patients’ motor proficiency and ensuring the adaptability of feedback systems to meet their specific needs. Existing literature supports the benefits of multimodal feedback in improving both motor performance and learning [39, 40, 41].The qualitative data in this study reinforce this perspective, demonstrating that individuals rely on different sensory inputs based on personal preference and prior experience. Some participants favored tactile cues for their immediacy and clarity, while others found auditory feedback more intuitive. Studies indicate that auditory input enhances tactile processing and goal-directed motor behavior, leading to faster and more accurate responses [42]. Additionally, the combination of auditory and tactile stimuli improves localization accuracy and accelerates reaction times, further emphasizing the role of cross-modal interactions in motor performance [43]. This principle of superadditivity, where multiple sensory inputs results in enhanced physiological responses beyond the sum of individual inputs, has been well-documented in neurophysiological research [44]. Moreover, multisensory interactions are particularly relevant in processing stimuli close to the body, where tactile and auditory cues naturally co-occur [45]. The superior colliculus, a key structure in sensory integration, plays a fundamental role in shortening response latencies when multimodal stimuli are present, highlighting the importance of combining sensory feedback for more effective motor behavior [44]. To conclude, the ability to integrate auditory and haptic information can enhance motor responses and body awareness. Future research should focus on tailoring feedback strategies based on skill level and determining the optimal timing and adaptation mechanisms to maximize both motor performance and learning outcomes in physiotherapeutic and motor training contexts.

### 4.4 Feedback usefulness

One significant result in this study concerns the higher number of repetitions given in the haptic condition compared to the auditory and combined feedback conditions. Several hypotheses could explain this finding:

1. Increased Cognitive Load for Haptic Feedback: Haptic feedback may initially be more difficult to interpret, requiring more repetitions for participants to adjust. This aligns with research indicating that haptic feedback can demand additional cognitive resources, particularly in unfamiliar tasks [46].

2. Temporal Constraints of Auditory Feedback: The auditory modality (and thus the combined) requires more time for feedback delivery due to the length of verbal instructions compared to the brevity of haptic vibrations. This suggests that fewer repetitions in the auditory and combined conditions may be due to practical constraints on feedback frequency, rather than differences in perceived usefulness.

3. Influence of Actuator Adherence and Body Fit: The effectiveness of haptic feedback may have been affected by variation in actuator contact with the skin, influenced by individual differences in body weight and garment fit. This is supported by findings that feedback perception can be modulated by tactile intensity and placement [47].

Despite the higher number of repetitions required for haptic feedback, qualitative data suggest that participants found haptic feedback more intuitive than auditory feedback. This aligns with previous findings that tactile cues can be processed more rapidly than auditory ones, leading to faster motor responses in some contexts [48]. However, participants also highlighted the value of combined feedback, particularly in making haptic cues easier and faster to interpret [47, 48]. This suggests that multimodal feedback strategies may provide a more accessible and adaptable user experience. These findings align with our prior work on feedback in physiotherapy and motor training, which emphasizes the importance of multimodal integration for effective motor learning [49]. Physical therapists often rely on auditory feedback or multimodal approaches that incorporate verbal instructions, revealing that different feedback modalities align with distinct task requirements. Visual or auditory feedback was linked to correcting the initial position. Whereas, tactile cues were associated with spatial movement and muscle activity. This suggests that future feedback design should tailor unimodal vs. multimodal strategies to specific motor learning goals, ensuring that each sensory channel optimally supports a given task.

### 4.5 Implications

The study overall proved the great potential of a multisensory feedback shirt to help people in physiotherapy. Despite the lack of statistically significant results, qualitative data analysis proved the perceived usefulness of the system in helping them perform the given exercise. As discussed in the Body Image Representation section, the brain integrates visual, tactile, and proprioceptive information to construct a coherent body image, suggesting that the effectiveness of feedback in physiotherapy is deeply rooted in multisensory integration. Additionally, research on motor performance improvement reinforces that multimodal feedback strategies facilitate faster and more efficient motor responses [44, 45]. These findings suggest that incorporating multiple sensory channels could enhance both the precision and efficiency of rehabilitation exercises, improving overall patient outcomes. An implicit result that seems to emerge from the qualitative study is the perceived advantage of the haptic feedback: This finding aligns with the growing recognition of social affective touch in rehabilitation contexts, where tactile stimuli can instill confidence and enhance engagement [50]. However, results from the Feedback Usefulness section suggest that haptic feedback is most effective when complemented by auditory input, particularly in the early stages of learning. This supports prior evidence that multimodal feedback optimizes information processing, motor learning, and user experience [47, 48]. The combination of sensory inputs, particularly the auditory and tactile cues, creates a more accessible and engaging experience for users, particularly for individuals who are less familiar with the motor task or feedback system. Our findings reinforce the idea that no single feedback modality is universally optimal. Instead, feedback perception is highly individual, influenced by sensory preferences, cognitive processing differences, and technical factors like actuator adherence and garment fit. As observed in the Performance Improvement section, different individuals prioritize different sensory channels, suggesting that a one-size-fits-all approach is insufficient. The most critical requirement for an effective multimodal feedback system is its ability to be tailored by a qualified expert to match the specific needs and clinical conditions of each patient. Personalized adjustments to the feedback system, such as varying the intensity or frequency of the sensory cues, would ensure that patients receive optimal support in their rehabilitation journey. Ultimately, our interdisciplinary approach—combining neuroscientific insights on body representation, sports science findings on motor learning, and user experience research on feedback perception—demonstrates that multimodal feedback is not only beneficial but necessary for designing adaptive, accessible, and effective rehabilitation technologies.

### 4.6 Limitations

This study has several limitations, both of a technical and statistical nature. The first limitation is that the same shirt was used for all 57 participants. This could, as said earlier, have impacted on the perception of the haptic feedback in some participants; clips were used for the participants on which the shirt was too large, but in some situations, the feedback was not perceived well enough. Furthermore, it should be noted that for the entire experiment, just one physiotherapy exercise was performed; this was done to avoid adding other variables to the study, but it is possible that some of the results are influenced by the chosen exercise. It is also important to note that the participants’ performance could have been influenced by the presence of the two experimenters during the study; some answers to the questionnaires or the performance of the exercise itself could have been influenced by social desirability or by a feeling of pressure. Another limitation of the study is connected to the high number of variables that were taken into consideration: the statistical values for some of the tests conducted were lowered by this. A final limit is the language of the study: the decision was taken to use both German and English-speaking participants, and despite a double-validated version for nearly each of the questionnaires used, for some, a modified version was created, but they are missing statistical validation.

### 4.7 Future studies

This study lays a strong foundation for developing a multisensory feedback system aimed at supporting individuals in physiotherapy. Future research should focus on quantitatively confirming the hypothesis that haptic feedback has a social, affective impact on patient engagement and motivation, as observed in the qualitative data. Additionally, further studies are needed to optimize the balance between sensory inputs and refine personalization strategies to maximize the effectiveness of multisensory feedback in physiotherapy exercises. Ensuring that these systems are adaptable to the diverse needs of patients is crucial for improving therapeutic outcomes.

## Data Availability

All data produced in the present study are available upon reasonable request to the authors.

## 5. Acknowledgements

Funded by Else Kröner Fresenius Center for Digital Health (EKFZ), University of Technology Dresden (TU Dresden), Dresden, Germany Funded by the German Research Foundation (DFG, Deutsche Forschungsgemeinschaft) as part of Germany’s Excellence Strategy-EXC 2050/1-Project ID 390696704-Cluster of Excellence “Centre for Tactile Internet with Human-in-the-Loop” (CeTI) of Technische Universität Dresden. The authors acknowledge the financial support by the Federal Ministry of Education and Research of Germany in the programme of “Souverän. Digital. Vernetzt.”. Joint project 6G-life, project identification number: 16KISK001K

## Declaration of generative AI and AI-assisted technologies in the writing process

During the preparation of this work the authors used Chat AI in order to improve language and readability, with caution. After using this tool, the authors reviewed and edited the content as needed and take full responsibility for the content of the publication.

## Appendix A. Supplementary

**Table A1.** Mean and standard deviation for participants’ age, weight and height. Note that 1 non-binary participant took part in the study as well; its data are included in the overall column.

|  | FEMALE | MALE | OVERALL |
| --- | --- | --- | --- |
| N | 29 | 27 | 57 |
| Mean age (years) | 28.83 | 28.78 | 28.88 |
| SD age (years) | 10.89 | 10.49 | 10.52 |
| Mean weight (kg) | 63.68 | 77.56 | 70.54 |
| SD weight (kg) | 8.04 | 11.18 | 11.79 |
| Mean height (cm) | 167.4 | 181.4 | 174.4 |
| SD height (cm) | 6.92 | 7.80 | 10.26 |

**Table A2.** Full output for the ANCOVA for the analysis of workload on participants in different modality conditions, controlling for users’ trust in technology (SRTQ and TIA questionnaires), as well as for demographic variables such as gender, age, height and weight. Significance codes as following: ‘**’ 0.01, ‘*’ 0.05

|  | Df | Sum Sq | Mean Sq | F Value | Pr (>F) |
| --- | --- | --- | --- | --- | --- |
| Feedback modality | 2 | 114 | 57.1 | 0.323 | 0.72468 |
| Characteristics and reliability (SRTQ) | 1 | 41 | 41 | 0.231 | 0.63123 |
| Perceived risk (SRTQ) | 1 | 27 | 27.1 | 0.153 | 0.69615 |
| Knowledge of the system (SRTQ) | 1 | 385 | 385.2 | 2.176 | 0.14232 |
| Reliability/Competence (TIA) | 1 | 480 | 480.3 | 2.714 | 0.10166 |
| Understanding/Predictability (TIA) | 1 | 336 | 335.5 | 1.896 | 0.1707 |
| Familiarity (TIA) | 1 | 940 | 940.2 | 5.312 | 0.0226* |
| Intention of Developers (TIA) | 1 | 1299 | 1298.5 | 7.336 | 0.00757** |
| Propensity to Trust (TIA) | 1 | 192 | 191.7 | 1.083 | 0.2998 |
| Trust in automation (TIA) | 1 | 1101 | 1100.5 | 6.218 | 0.01377* |
| Age | 1 | 405 | 404.7 | 2.286 | 0.13268 |
| Height | 1 | 1433 | 1432.8 | 8.095 | 0.00508** |
| Weight | 1 | 230 | 229.6 | 1.297 | 0.25662 |
| Gender | 2 | 177 | 88.7 | 0.501 | 0.60684 |
| Residuals | 145 | 25665 | 177 |  |  |

**Table A3.** Full output for the ANOVA for the analysis of body image representation in different modality conditions.

|  | Df | Sum Sq | Mean Sq | F Value | Pr |
| --- | --- | --- | --- | --- | --- |
| Feedback modality | 2 | 3 | 1.69 | 0.012 | 0.988 |
| Residuals | 681 | 94308 | 138.48 |  |  |

**Table A4.** Full output for the ANOVA for the analysis of the improvement in the performance in different feedback modalities, measured by the first three items of the Physiotherapist evaluation form for correction behaviour on 4-point Likert scale.

|  |  | Df | Sum Sq | Mean Sq | F Value | Pr |
| --- | --- | --- | --- | --- | --- | --- |
| Item 1 | Feedback modality | 2 | 0.29 | 0.1462 | 0.579 | 0.561 |
|  | Residuals | 168 | 42.39 | 0.2523 |  |  |
| Item 2 | Feedback modality | 2 | 1.56 | 0.7778 | 1.315 | 0.271 |
|  | Residuals | 168 | 99.40 | 0.5917 |  |  |
| Item 3 | Feedback modality | 2 | 0.38 | 0.1914 | 0.944 | 0.391 |
|  | Residuals | 159 | 32.24 | 0.2028 |  |  |

**Table A5.** Full output for the ANCOVA for the analysis of number of feedback in different modality conditions, controlling for number of repetitions, item 4 of the Physiotherapist evaluation form (Reaction Speed), and users’ trust in technology (SRTQ and TIA questionnaires), as well as for demographic variables such as gender, age, height and weight. Significance codes as following: ‘***’ 0.001, ‘**’ 0.01, ‘*’ 0.05.

|  | Df | Sum Sq | Mean Sq | F Value | Pr |
| --- | --- | --- | --- | --- | --- |
| Feedback modality | 2 | 12.05 | 6.025 | 8.333 | 0.000419*** |
| Number of repetitions | 1 | 9.4 | 9.396 | 12.996 | 0.000464*** |
| Reaction speed | 1 | 20.13 | 20.132 | 27.846 | 6.34E-07*** |
| Characteristics and reliability (SRTQ) | 1 | 2.28 | 2.281 | 3.154 | 0.078392 |
| Perceived risk (SRTQ) | 1 | 2.16 | 2.164 | 2.993 | 0.086339 |
| Knowledge of the system (SRTQ) | 1 | 0.16 | 0.163 | 0.226 | 0.635735 |
| Reliability/Competence (TIA) | 1 | 0.62 | 0.624 | 0.863 | 0.354759 |
| Understanding/Predictability (TIA) | 1 | 1.01 | 1.014 | 1.403 | 0.238704 |
| Familiarity (TIA) | 1 | 0.94 | 0.94 | 1.3 | 0.256643 |
| Intention of Developers (TIA) | 1 | 0.65 | 0.646 | 0.893 | 0.34669 |
| Propensity to Trust (TIA) | 1 | 1.94 | 1.938 | 2.68 | 0.104365 |
| Trust in automation (TIA) | 1 | 0.78 | 0.784 | 1.085 | 0.299821* |
| Age | 1 | 4.23 | 4.232 | 5.853 | 0.017132 |
| Height | 1 | 0 | 0.005 | 0.007 | 0.934654 |
| Weight | 30 | 42.44 | 1.415 | 1.957 | 0.006165** |
| Gender (Male/Female/Non-binary) | 2 | 1.78 | 0.892 | 1.233 | 0.295185 |
| Residuals | 114 | 82.42 | 0.723 |  |  |

**Table A6.** Post-hoc comparison conducted using the t Test with Bonferroni correction for number of feedbacks in different modalities. ‘*’ indicates a p-value lower than alpha corrected with Bonferroni correction (0.0167).

|  | t | df | mean difference | 95% CI (lower) | 95% CI (upper) | p-value |
| --- | --- | --- | --- | --- | --- | --- |
| Haptic-Auditory | 3.988 | 53 | 0.667 | 0.331 | 1.002 | 0.0002058* |
| Haptic-Combined | 2.358 | 53 | 0.370 | 0.055 | 0.685 | 0.02208 |
| Combined-Auditory | 1.993 | 53 | 0.296 | -0.002 | 0.594 | 0.05143 |

## Notes

### Competing Interest Statement

The authors have declared no competing interest.

### Funding Statement

Funded by Else Kroner Fresenius Center for Digital Health (EKFZ), University of Technology Dresden (TU
Dresden), Dresden, Germany Funded by the German Research Foundation (DFG, Deutsche Forschungsgemeinschaft) as part of Germanys Excellence Strategy-EXC 2050/1-Project ID 390696704-Cluster of Excellence Centre for Tactile Internet with Human-in-the-Loop (CeTI) of Technische Universitat Dresden. The authors acknowledge the financial support by the Federal Ministry of Education and Research of Germany in the programme of Souveran. Digital. Vernetzt.. Joint project 6G-life, project identification number: 16KISK001K.

### Author Declarations

The research project was approved by the Ethics Committee of Technische Universitat Dresden (BOEK- 215052022) and follows the Declaration of Helsinki.

